# Are Vapers More Susceptible to COVID-19 Infection?

**DOI:** 10.1101/2020.05.05.20092379

**Authors:** Dongmei Li, Daniel P. Croft, Deborah J. Ossip, Zidian Xie

## Abstract

**Background:** COVID-19, caused by severe acute respiratory syndrome coronavirus 2 (SARS-CoV-2), was declared a global pandemic in March 2020. Electronic cigarette use (vaping) rapidly gained popularity in the US in recent years. Whether electronic cigarette users (vapers) are more susceptible to COVID-19 infection is unknown.

**Methods:** Using integrated data in each US state from the 2018 Behavioral Risk Factor Surveillance System (BRFSS), United States Census Bureau and the 1Point3Acres.com website, generalized estimating equation (GEE) models with negative binomial distribution assumption and log link functions were used to examine the association of weighted proportions of vapers with number of COVID-19 infections and deaths in the US.

**Results:** The weighted proportion of vapers who used e-cigarettes every day or some days ranged from 2.86% to 6.42% for US states. Statistically significant associations were observed between the weighted proportion of vapers and number of COVID-19 infected cases as well as COVID-19 deaths in the US after adjusting for the weighted proportion of smokers and other significant covariates in the GEE models. With every one percent increase in weighted proportion of vapers in each state, the number of COVID-19 infected cases increase by 0.3139 (95% CI: 0.0554 –0.5723) and the number of COVID-19 deaths increase by 0.3705 (95% CI: 0.0623 – 0.6786) in log scale in each US state.

**Conclusions:** The positive associations between the proportion of vapers and the number of COVID-19 infected cases and deaths in each US state suggest an increased susceptibility of vapers to COVID-19 infections and deaths.

## Introduction

Novel coronavirus disease 2019 (COVID-19) outbreak was declared a global pandemic by the World Health Organization (WHO) on March 11, 2020. ^1^ As of April 28, 2020, there were over three million COVID-19 infected cases and over 200,000 deaths globally. ^2^ In the United State, the total number of infected COVID-19 cases exceeded one million, with over 57,000 deaths reported by April 28, 2020. COVID-19 infection presents with cough, dyspnea and fever among other systemic symptoms and can lead to pneumonia and acute hypoxemic respiratory failure. ^3^

Electronic cigarettes (e-cigarettes), promoted as an alternative for cigarette smoking, rapidly gained popularity in recent years in the US. In 2018, the prevalence of current e-cigarette use (vaping) in US adults was 3.2%. ^4^ Recent studies on the associations of vaping and health symptoms/diseases have observed associations between vaping and symptoms of wheezing and self-reported Chronic Obstructive Pulmonary Disease (COPD), along with increased inflammation in bronchial epithelial cells and alterations in the pulmonary immune response to infection. ^5-10^ Tobacco control researchers have raised concerns that vapers may be more susceptible to COVID-19 infections and could develop more severe COVID-19 symptoms. ^11^ However, there is very limited evidence on the association between vaping and COVID-19 infection.

We will examine the association of vaping with COVID-19 infections and deaths, using the integrated state-level weighted proportions of current e-cigarette users (vapers) from the 2018 Behavioral Risk Factor Surveillance System (BRFSS) survey data, the population size and land area in 2018 in each state from United States Census Bureau, and the daily number of COVID-19 infected cases and deaths in each state from the 1Point3Acres.com website during the time period from January 21, 2020 to April 25, 2020 in the United States. Our study is the first one to provide evidence on the association of vaping with COVID-19 infections and deaths at the US population level.

## Methods

### Study Population

We integrated the 2018 Behavioral Risk Factor Surveillance System (BRFSS) survey data at state level, the population size and land area in each state from the United States Census Bureau, and the COVID-19 infected cases and deaths data from the 1Point3Acres.com website at available dates from each state through the unique two letter state abbreviations. From the 2018 BRFSS survey, 34 states provided information on the vaping status variable. The population size in each state in 2018 and land area in each state were obtained from the United States Census Bureau website. The COVID-19 infected cases and deaths counts were available for each state from January 21, 2020 to April 25, 2020. Reports of negative numbers of infected cases and deaths were excluded from the COVID-19 data. After integrating the BRFSS data and the census data with the COVID-19 infected case and deaths from different dates at the state level, there were 1607 observations in the final analysis data.

### Vaping Status

The vaping status variable was defined by the answers to the question “Do you now use e-cigarettes, every day, some days, or not at all?” in the 2018 BRFSS survey. Subjects who now use e-cigarettes every day or some days were classified as vapers and subjects who responded that they use e-cigarettes “not at all” or “not applicable” were classified as non-vapers. The weighted frequency of vapers in each state was obtained using the proc surveyfreq procedure in SAS version 9.4 (SAS Institute Inc., Cary, NC), considering the complex sampling design of the BRFSS survey. The weighted proportion of vapers in each US state was calculated using the ratio of weighted frequency of vapers and weighted frequency of total number of subjects in each state.

### Outcomes and Covariates

The outcomes used in current analysis are the number of COVID-19 infected cases and deaths. Covariates considered in the current investigation include population size, population density (calculated using population size divided by land area), age, gender, race/ethnicity, education, income, mental health, physical health, obesity, respiratory disease (including asthma and COPD), heart disease, cancer, stroke, diabetes, kidney disease, and smoking (currently smoke every day or some days). The number of COVID-19 infected cases was also used as a covariate when modeling the COVID-19 deaths.

### Statistical Analysis

Generalized estimating equation (GEE) models with negative binomial distribution assumptions and log link functions were used to examine the association of weighted proportion of vapers with the number of COVID-19 infections and deaths, after adjusting for the confounding effects from significant covariates. ^12,13^ The correlations of number of COVID-19 infections and deaths from different dates within the same state were considered through the autoregressive 1 (AR (1)) variance-covariance structure within the GEE model framework. The purposeful covariates selection method was used to select significant covariates for the GEE models. ^14^ Variance inflation factor (VIF) was used to examine the multicollinearities among the predictor variables in the GEE models. ^15^ A VIF value of five or less was considered to indicate multicollinearity in the fitted GEE model. All statistical analyses were conducted using statistical analysis software SAS version 9.4 (SAS Institute Inc., Cary, NC) and R (R Core Team, 2017). Significance levels for all tests were set at 5% for two-sided tests.

## Results

The weighted proportion of vapers ranged from 2.86% to 6.42% for US states. The daily number of infected COVID-19 cases ranged from 0 to 11,743 with an average of 362 daily cases across all states in the US during the time period from January 21, 2002 to April 25, 2020. During the same time period, the daily number of deaths ranged from 0 to 4,556 with the mean number of daily deaths at 20 across all US states.

Figure 1 shows the estimated coefficients and their 95% confidence intervals from the GEE model on daily number of COVID-19 infected cases. States that had a higher proportion of vapers also had a larger number of daily COVID-19 infected cases. For every one percent increase in the proportion of vapers, the log number of daily COVID-19 infected cases increased by 0.3139 (95% CI: 0.0554 – 0.5723) on average. The covariates included in the GEE model were state population size in log scale, male, less than high school education, poor physical health, cancer, obese, and smoker. Both population size in log scale in each state and proportion of less than high school education had a positive association with the daily COVID-19 infection. Male gender, poor physical health, cancer, and obesity had a negative association with the daily COVID-19 infection. Proportion of current smokers in each state was not significantly associated with the daily COVID-19 infected cases in each state.

**Figure 1.**
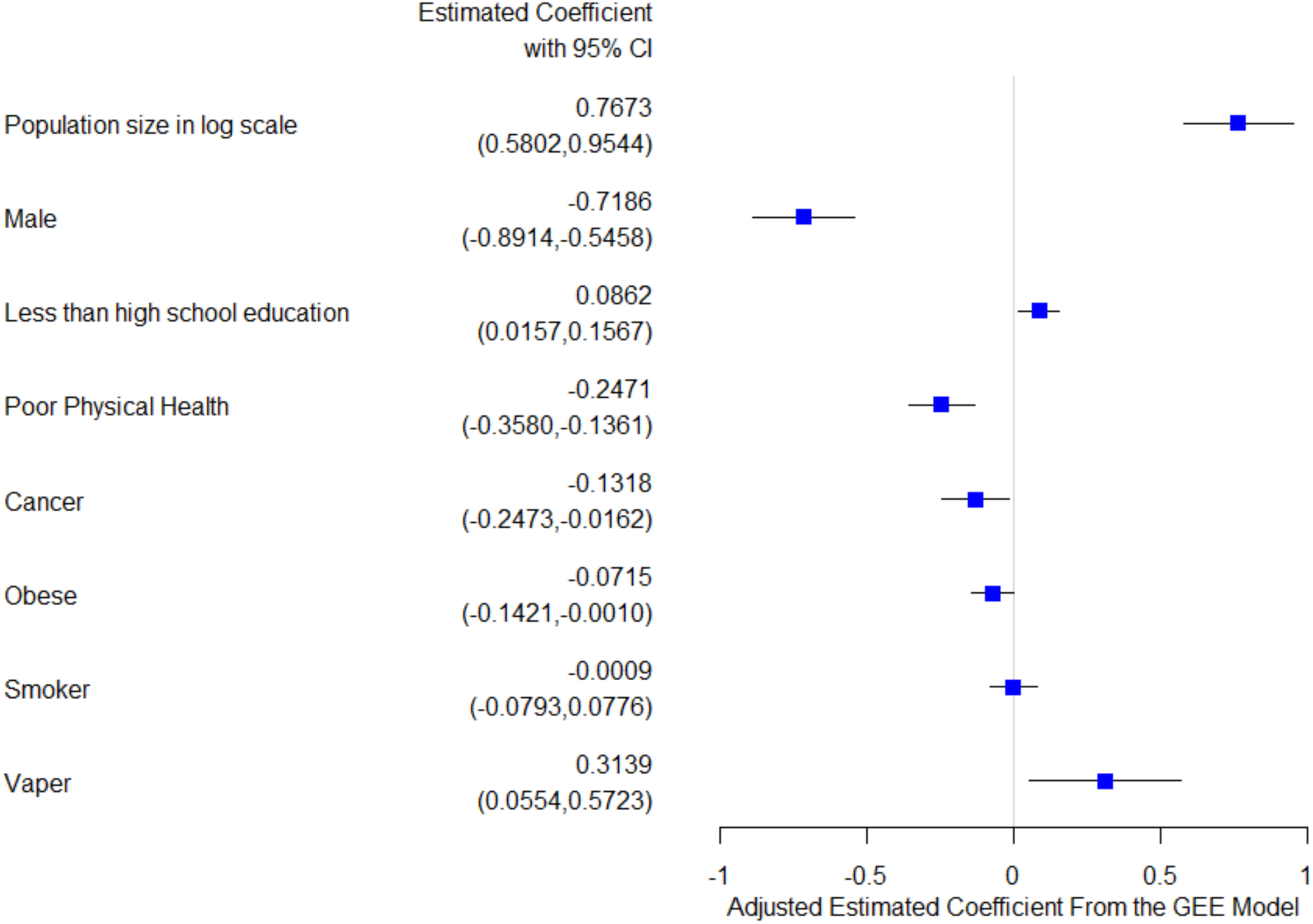
Adjusted estimated coefficients from the GEE model on number of COVID-19 infected cases.

The estimated coefficients and their 95% confidence intervals from the GEE model on daily number of COVID-19 deaths were shown in Figure 2. States that had a higher proportion of vapers also had a higher number of daily COVID-19 deaths. With a one percent increase in the proportion of vapers, the log number of COVID-19 deaths will increase 0.3705 (95% CI: 0.0623 – 0.6786) on average. The covariates in the GEE model included the number of daily confirmed cases in each state, state population size in log scale, proportion of people aged 35-44, proportion of White, Black, and Hispanic, proportion of people having respiratory disease, cancer, being obese, and being smokers. Daily confirmed cases, state population size in log scale, proportion of White, Black, and Hispanic, and proportion of people having respiratory disease were positively associated with the daily number of COVID-19 deaths in each state. The proportion of people aged 35-44, having cancer, and being obese all were negatively associated with daily number of COVID-19 deaths in each state. Similarly, the proportion of current smokers in each state was not significantly associated with the daily number of COVID-19 deaths in each state.

**Figure 2.**
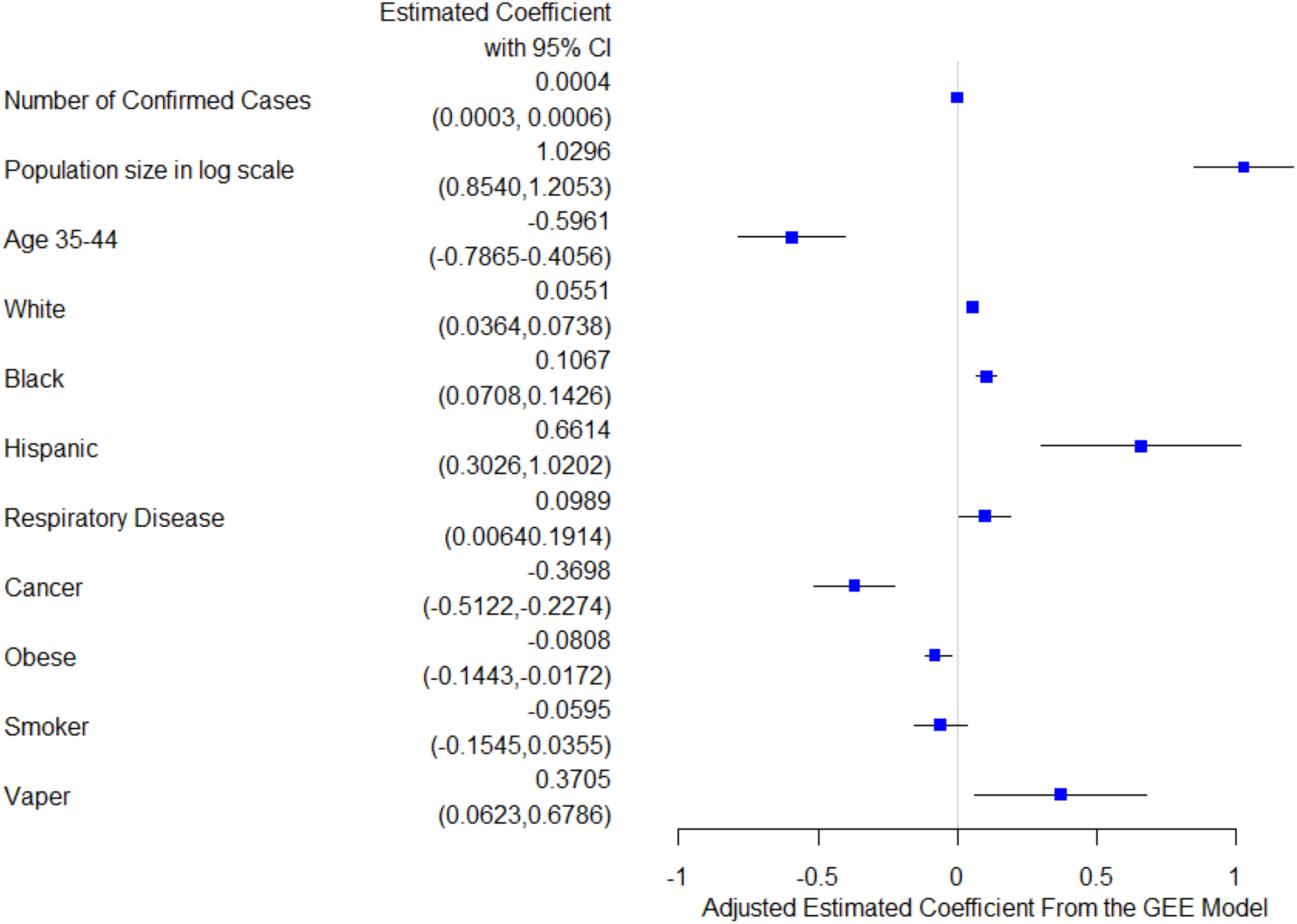
Adjusted estimated coefficients from the GEE model on number of COVID-19 deaths.

## Discussion

Using integrated state level data obtained from the 2018 BRFSS survey, the United States Census Bureau and the 1Point3Acres.com website, we were able to investigate the association of weighted proportion of vapers with COVID-19 infected cases and deaths in the United States. We found a significant positive association of vaping with COVID-19 infections and deaths at the state level controlling for sociodemographic and health related covariates. This finding further supports future research into the potential susceptibility of vapers to COVID-19 infection. Additional findings include the expected positive association between population size and COVID-19 infection and death. Finally we also observed a positive association between a low education level and COVID-19 infection.

Though we identified a positive association between the weighted proportion of vapers and the number of COVID-19 infections and deaths at state level, we did not have data on what proportion of those who actually contracted COVID-19 or died from COVID-19 were vapers. Thus, we cannot determine causality between vaping and COVID-19 infections and deaths. However, prior research supports the biological plausibility of a relationship between vaping and an increased susceptibility to respiratory infection. ^16^ Multiple mouse models have observed an increased severity in infection associated with vaping exposure related to dysregulation of lung epithelial cells and an impaired immune response to both viral^17^, and bacterial infection. ^18^ Bacterial superinfection of viral illnesses like influenza and COVID-19 is especially dangerous, as this leads to an increased severity in illness ^19^ A human cell based model of exposure to nicotine-free flavored e-liquid observed immunosuppressive effects and impaired respiratory innate immune cell function (alveolar macrophages, neutrophils, and natural killer cells).^20^ In humans, bronchoalveolar lavage samples from the airways of active vapers also revealed dysregulation of the airway’s innate immune response including neutrophilic response and mucin ^21^ A wide variety of flavorings are used by vapers, many of which, such as diacetyl, acetoin, pentanedione, o-vanillin, maltol, and coumarin in nicotine-free e-liquid, could also trigger inflammatory responses in human monocytes.^7^ Finally, previous human studies found vaping is associated with increased risk of chronic bronchitic symptoms (chronic cough, phlegm, or bronchitis).^22-24^ and epidemiologic studies observed an increased risk of self-reported wheezing and COPD. ^9,10^ With the ongoing COVID-19 pandemic, particular health concerns have been raised regarding vaping, such as whether vapers have higher risk for COVID-19 infection and could develop more severe symptoms once contracted COVID-19.^11^ To our best knowledge, this is the first population-based study to empirically examine and find an association between vaping and COVID-19.

The existing literature on the increased risk of respiratory infection in combustible cigarette smokers was summarized by a prior meta-analysis finding an increased risk of current smokers for influenza infection compared to non-smokers.^25^ As COVID-19 is a novel condition, the literature examining the risk of smokers for COVID-19 is scant. A recent study based on 1099 COVID-19 patients found smoking history was associated with COVID-19 severity.^26^ A recent systematic review on COVID-19 and smoking concludes that smoking is likely associated with worse outcomes in COVID-19.^27^ However, other studies indicate that smoking might not be associated with the incidence and severity of COVID-19 ^28^, for example, a recent meta-analysis based on Chinese patients suggests that active smoking is not associated with severity of COVID-19.^29^ It remains unclear whether nicotine has a role in the either the increased or decreased severity of illness for smokers with COVID-19. Our study did not find a significant association between the weighted proportion of smokers and the number of COVID-19 infections and deaths at state level. Due to the incomplete testing and tracking of home deaths, it is possible that a percentage of older smokers with comorbidities are dying at home from COVID-19 and therefore are not captured into the reported COVID-19 infections and death data ^30,31^ Another possibility is that smokers with comorbidities are homebound and more likely to be strictly following social/physical distancing guidelines, and are therefore reducing their risk for COVID-19. Currently, there is limited evidence on the susceptibility of smokers to COVID-19 infection and whether smokers have a worsened course in the setting of COVID-19. Additionally, it is unknown whether or not COVID-19 virus could be transmitted to those surrounding smokers through passive smoking and vaping. More epidemiological and clinical studies are needed to investigate the association of smoking with COVID-19 infections and deaths.

We found states that had a larger weighted proportion of subjects who had less than high school education had a higher number of COVID-19 infections. Researchers from the University of Southern California found that Americans who had less than high school education had a lower perceived risk of exposure to COVID-19 and a higher perceived risk of deaths than those who have college or higher degrees. ^32^ This might explain, in part, the positive association between the weighted proportion of less than high school education with the number of COVID-19 infections. We also found states that had a larger proportion of non-Hispanic Blacks and Hispanics had a larger number of COVID-19 deaths. The COVID-19 deaths rate data from Washington D.C. and 36 US states reported through April 27, 2020 showed that non-Hispanic Blacks (28.4%) and Hispanics (11.3%) had the highest COVID-19 deaths rate. ^33^ This could be related to the higher proportion of chronic conditions such as hypertension, heart disease and diabetes in non-Hispanic Blacks and Hispanics.^34,35^ We also found that states having a higher proportion of respiratory disease such as asthma or COPD had a higher number of COVID-19 deaths, which indicated that respiratory disease such as asthma and COPD could potentially increase the risk of COVID-19 deaths.

There are several limitations in current study. One limitation is that the weighted proportions of vapers, smokers, and other demographic and chronic diseases are from the 2018 BRFSS data, which might differ from the 2020 estimates. The reported COVID-19 infected cases and deaths obtained from 1Point3Acres.com website could be subject to some reporting errors as we noticed some negative number of COVID-19 infected cases and deaths, which we excluded from further analysis. However, we compared the COVID-19 data obtained from 1Point3Acres.com website with the COVID-19 data from the New York Times ^36^ and Centers for Disease Control and Prevention (CDC) website ^37^ and found consistent numbers of COVID-19 infections and deaths, which increased the robustness and reliability of the data sources. Another limitation is that our current analysis is on state level instead of individual level. We don’t know the individual status of vaping and COVID-19 infections or deaths, thus estimated coefficients of association could be different from epidemiological or clinical studies on individual subjects.

## Conclusions

There are positive associations between the weighted proportion of vapers in each US state and the daily number of COVID-19 infections and deaths in each US state. These positive associations suggest vapers may have an increased susceptibility to COVID-19 infections and deaths. Systematic assessment of vaping among patients, along with additional studies on the associations of vaping with COVID-19 infections and deaths at individual level are needed to confirm the identified positive association between vaping and COVID-19 infections and deaths.

## Data Availability

The 2018 Behavioral Risk Factor Surveillance System (BRFSS) survey data are publicly available from the Centers for Disease Control and Prevention website (https://www.cdc.gov/brfss/annual_data/annual_2018.html). The state population in 2018 and the land area in each state were obtained from the United States Census Bureau website (https://www.census.gov/). The COVID-19 infected cases and deaths data were requested and obtained from the 1Point3Acres.com website (https://coronavirus.1point3acres.com/en).

https://www.census.gov/

https://coronavirus.1point3acres.com/en

## Contributors

DL, ZX: conceived and designed the study. DL: analyzed the data. DL wrote the manuscript.

DL, DC, DO, ZX: assisted with interpretation of analyses and edited the manuscript.

## Disclaimer

The content is solely the responsibility of the authors and does not necessarily represent the official views of the NIH or the FDA.

## What this paper adds

- Current e-cigarette use is positively associated with COVID-19 infections.
- Current e-cigarette use is positively associated with COVID-19 deaths.
- This study emphasizes the importance of studying the susceptibility of current e-cigarette users to COVID-19 infection and death.

